# Neural Correlates and Functional Cognitive Maps in Breast Cancer Survivors Receiving Different Chemotherapy Regimens; a QEEG/HEG – based Investigation

**DOI:** 10.1101/2021.08.28.21262758

**Authors:** Maryam Vasaghi Gharamaleki, Seyedeh Zahra Mousavi, Maryam Owrangi, Mohammad Javad Gholamzadeh, Ali-Mohammad Kamali, Mehdi Dehghani, Prasun Chakrabarti, Mohammad Nami

## Abstract

**Background:** Post-chemotherapy cognitive impairment commonly known as “chemobrain” or “chemofog” is a well-established clinical disorder affecting various cognitive domains including attention, visuospatial working memory, executive function, etc. Although several studies have confirmed the chemobrain in recent years, scant experiments have evaluated the potential neurotoxicity of different chemotherapy regimens and agents. In this study, we aimed to evaluate the extent of attention deficits, one of the commonly affected cognitive domains, among breast cancer patients treated with different chemotherapy regimens through neuroimaging techniques.

**Methods:** Breast cancer patients treated with two commonly prescribed chemotherapy regimens, AC-T and TAC, and healthy volunteers were recruited. Near-infrared hemoencephalography (nirHEG) and quantitative electroencephalography (qEEG) assessments were recorded for each participant at rest and during task performance to compare the functional cortical changes associated with each chemotherapy regimen.

**Results:** The qEEG analysis revealed increased power of high alpha/low beta or sensorimotor rhythm (SMR) frequency in left fronto-centro-parietal regions involved in dorsal and ventral attention networks (DAN and VAN) in the AC-T-treated group comparing to the TAC and control group. The AC-T group also had the highest current source density (CSD) values in DAN and VAN-related centers in 10 and 15 Hz associated with the lowest Z-scored FFT coherence in the mentioned regions.

**Conclusions:** The mentioned findings revealed increased cognitive workload and lack of cognitive ease in breast cancer patients treated with the AC-T regimen proposing the presumable neurotoxic sequelae of this chemotherapy regimen in comparison with the TAC regimen.

## 1. Introduction

Breast cancer is currently the most commonly diagnosed cancer and the 5th cause of cancer deaths worldwide. According to “GLOBOCAN” estimates, it accounts for about 24.5% of new cancer cases and 15.5% of cancer deaths in 2020 among females (1, 2). It seems that both premenopausal and postmenopausal incidences of breast cancer are increasing worldwide due to lifestyle and hormonal risk factors and increased screening rates (3). Since the improvement of diagnostic and treatment strategies has increased the breast cancer survival rate, improving survivors’ quality of life (QoL) is of great importance (4). A study conducted by Schmidt et al. revealed that although QoL in breast cancer survivors substantially improved over time, many survivors still suffered from post-chemotherapy complications with cognitive impairment as the most disabling problem (5). Cognitive performance is one of the determinants of Qol which impairment can persist for several years in breast cancer survivors (6-9). As chemotherapy is the mainstay of breast cancer treatment, its side effects especially its neurotoxic sequelae should be considered to make less impact on the patient’s QoL.

The chemotherapy-induced cognitive impairment, commonly known as “chemobrain” or “chemofog”, has been reported among breast cancer survivors subjectively (10-14). Multiple studies have evaluated chemobrain prevalence in breast cancer survivors suggesting that the chemobrain incidence rate can be up to 70% (13, 15). However, these complaints have been reported more subjectively than objectively (16-18). One explanation is that chemotherapy is associated with a subtle cognitive impairment that can be latent in neuropsychological tests though problematic in patients’ real life (19).

A meta-analysis published by Ono et al. evaluated the performance of breast cancer patients who underwent adjuvant chemotherapy in eight cognitive domains and revealed that chemotherapy has a small but significant impact on cognitive function. Although the study showed that five out of eight domains of cognitive functioning can be influenced, it seems that some cognitive domains including processing speed can be more affected by chemotherapy and some domains such as long-term memory can improve over time (20).

Besides, numerous studies have attempted to assess chemobrain objectively mostly through neuroimaging assessments which were indicative of morphological changes including reduced gray matter in frontal, temporal, and parietal regions in cancer patients which correlates with cognitive decline (8, 21-25). In one longitudinal study accomplished by Deprez et al. using magnetic resonance diffusion tensor imaging (DTI), changes in cerebral white matter integrity in important tracts involved in cognition were demonstrated after chemotherapy in breast cancer patients (26). Also, another large cross-sectional study illustrated a long-term reduction in total brain volume and gray matter in cancer patients who received adjuvant chemotherapy via structural MRI (27). Moreover, applying fMRI has shown alterations in brain activation patterns particularly in the frontal lobe during tasks involving cognitive domains such as working memory in the chemotherapy-exposed patients (28). Nevertheless, limited studies have evaluated the impact of chemotherapy on functional brain network alterations by electroencephalography (EEG) (29).

On the other hand, Various chemotherapy regimens are used as adjuvant therapy to reduce mortality and recurrence in HER2-negative breast cancer. Selection of the appropriate regimen should be individualized regarding the patient’s characteristics. However, adverse effects and long-term complications of each regimen should be exactly monitored to help physicians prescribing the less toxic regimen. For most patients in whom chemotherapy is indicated, the selected regimen is dose-dense doxorubicin and cyclophosphamide (AC) followed by paclitaxel (T), known as the AC-T regimen. Another alternative is the TAC regimen in which docetaxel plus doxorubicin and cyclophosphamide are administered (30, 31). Despite vast research assessing the relationship between chemotherapy and cognitive decline, scant studies have compared neurotoxic effects of common chemotherapy regimens.

We have previously conducted a study evaluating the magnitude of cognitive impairment in chemotherapy-treated breast cancer patients through Addenbrooke’s Cognitive Examination (ACE) and Cambridge Brain Sciences (CBS) cognitive tasks. The study has revealed that the AC-T regimen can contribute to worse performance than the TAC regimen in cognitive tasks of monkey ladder and double trouble in which visuospatial working memory and response inhibition -aspects of memory and concentration- are involved (https://www.cambridgebrainsciences.com) (32).

Response inhibition, the multidimensional cognitive process of suppression of inappropriate action for coordination of desired tasks, provokes various neural systems mainly the fronto-parietal and ventral attention network (VAN) (33). Also, attentional allocation depends on the coordination between the ventral attention network (VAN) and dorsal attention network (DAN). The DAN comprises the dorsolateral prefrontal cortex (dlPFC), frontal eye fields, middle temporal motion complex, and superior parietal lobe, modulates the top-down attentional control (34). VAN, the other network involved in sustained attention, also comprises dorsomedial, mid and ventrolateral prefrontal cortex (PFC), anterior insula and parietal regions including intraparietal sulcus (IPS) and temporoparietal joint (TPJ), as well as midbrain, thalamus, basal ganglia, and cerebellum (35). Both the DAN and VAN can be stimulated simultaneously by exogenous stimulants (34).

Therefore, as response inhibition, visuospatial working memory, and sustained attention have shown a significant decline in chemotherapy-treated patients which provoke neuronal activation in DAN and VAN (33-36), we aim to assess the functional effects of the chemotherapy regimens mentioned above on these brain networks via two less commonly applied methods, electroencephalography and hemoencephalography (HEG), to determine the regimen with less neurotoxic sequela and help clinicians to make a more informed decision.

## 2. Methods and Materials

### 2.1 Patient Selection

The patients and control group enrolled in our previous study were asked to participate in the current experiment. Patients were recruited from Motahari Polyclinic, the affiliated medical center of Shiraz University of Medical Sciences, between 2018 and 2020. Participants were chemotherapy-treated breast cancer patients who met the following criteria: 1) women aged 30-65 years, 2) diagnosis of breast cancer was confirmed with trucut biopsy, 3) at least six months have been passed from their last chemotherapy cycle, and 4) all patients have at least a high school degree to perform cognitive tests and answer questionnaires. Exclusion criteria included: 1) presence of other neuropsychological comorbidities such as epilepsy, schizophrenia, bipolar disorder, depression, etc. that can substantially affect cognitive function, 2) brain metastasis, and 3) stage IV of the disease. The patients were furtherly divided into two groups based on the chemotherapy regimen.

The control group included women aged 30-65 years without a history of known malignancy or neuropsychological disorders that have been matched with the patient group in terms of age, education, and socio-economic status.

The Ethics Committee of Shiraz University of Medical Sciences, Shiraz, Iran has approved the experiment (IR.SUMS.REC.1398.303). All experiments were performed in accordance with relevant guidelines and regulations. All participants were informed of the purpose of the study and performed interventions and completed the informed consent before recruiting into the study.

### 2.2 Demographic information

All participants were asked to complete a demographic questionnaire including information regarding the age, marital status, education, occupation, past medical and drug history and in particular neuropsychiatric disorders, and detailed information of the underwent cancer treatments for breast cancer patients (e.g. chemotherapy regimen, radiotherapy, surgery, etc.).

### 2.3 Hemoencephalography

Near-infrared hemoencephalography (nirHEG), a neuroimaging method of monitoring changes in cortical blood circulation and oxygenation (37), was used in this study to measure prefrontal cortical hemodynamic changes. The NeXus-4 biofeedback and neurofeedback system (NeXus-4, Mind Media BV, Herten, the Netherlands) with a HEG sensor headband were utilized. The nirHEG headband has two optodes placed in contact with the skin, a light-generating LED pair for red and infrared light production, and a sensor optode to receive the returning light.

The nirHEG data were recorded from all participants at the left frontoparietal (FP1) area with the following method: 1) five minutes in resting-state sitting in a comfortable recliner with eyes open, 2) while performing one of the CBS cognitive tasks with the lowest Z score for each participant identified as the area needing improvement (ANI) according to the results of our previous study, and 3) five minutes post-test resting state with eyes open. BioTrace+ software (NeXus-4, Mind Media BV, Herten, the Netherlands) was used to analyze the HEG data.

### 2.4 Quantitative Electroencephalography (qEEG)

#### 2.4.1 qEEG acquisition

All participants underwent quantitative electroencephalography (qEEG) assessment using a 19-electrode EEG cap, fitted for each patient. The International 10-20 system was applied for electrode placement using a Linked Ears (LE) montage. EEG data were recorded using “NrSign EEG 3840” (NrSign Inc., Vancouver, Canada) with the sensitivity of 70 microvolts, a sampling rate of 500 samples/second, a low-cut filter at 0.5 Hz, and a high-cut filter at 70 Hz according to the following protocol: (1) five minutes in resting-state sitting in a comfortable recliner with eyes open, and then (2) while performing the ANI task.

#### 2.4.2 qEEG pre-processing

The raw EEG data was exported to “EEG Studio Processing” software (Mitsar Co. Ltd. St. Petersburg Russia) for pre-processing. Independent component analysis (ICA), a data decomposition method (38), was applied to extract artifacts from neuronal activity. ICA components related to eye blink and eye movement artifacts were removed. The pre-processed EEG data was then exported as the EDF file to “NeuroGuide” software version 3.0.9 (Applied Neuroscience, Inc., St. Petersburg, FL, USA), a comprehensive digital qEEG analysis system, for advanced processing.

To confirm the selection of artifact-free qEEG segments, automatic editing was also performed through NeuroGuide software. A 10-second artifact-free segment was manually selected by an expert in qEEG analysis as the template. The artifact-free template matching tool of the NeuroGuide software with defaults of high sensitivity for drowsiness and eye movement artifact rejection and amplitude multiplier of 1.00 was then applied to perform automatic editing. The artifact-free EEG selections with test re-test reliability > 0.90 and split-half reliability > 0.95 were confirmed for further analysis.

#### 2.4.3 qEEG analysis

To display and analyze functional cognitive maps of AC-T, TAC, and control groups separately, the edited EEG files of the resting-state of members of each group were combined to create a single text file. This text file was also imported to Neuroguide software and analyzed to produce cortical color topographic maps and other metrics including peak frequency, absolute power, relative power, coherence, phase lag, etc. representing the overall cognitive function of members of each group.

Besides, to enable the comparison, the edited artifact-free EEG selections of the resting-state of all the participants were converted to Neuroguide Analysis files (NGA) through NeuroGuide software that are of interest for comparison purposes. The Neurobatch toolbox was then applied to combine NGA files of all the members of a group and create Neuroguide Group analysis files (NGG) for the AC-T, TAC, and control groups separately. The three NGG files were then compared through group statistics analysis.

Neuronavigator analysis was also performed to assess the current source density (CSD) values, the volume density of net transmembrane currents (39), in DAN and VAN networks. Mean Frequencies of 10 and 15 Hz were selected to represent the high alpha and beta 1 frequency bands respectively. The Brodmann areas with the maximum CSD values in the mentioned frequencies in DAN and VAN networks among the control group were identified and selected as regions of interest (ROIs). The CSD values of these ROIs were then compared between the three groups. All the mentioned steps were furtherly repeated for analyzing the qEEG files of ANI task performance.

## 3. Results

### 3.1 Demographic information

In total, 24 healthy volunteers and 35 chemotherapy-treated breast cancer patients that all have been enrolled in our previous study accepted to participate in the current experiment, of whom 16 patients had received the AC-T chemotherapy regimen and 19 patients had been treated with the TAC regimen. All of the patients had a biopsy report confirming the breast malignancy pathologically, negative history of brain metastasis, and underwent a total or partial mastectomy, radiotherapy, and chemotherapy. Also, 24 healthy volunteers matched with the breast cancer patients in terms of age and education enrolled in the study as the control group.

The mean ages of the participants were 48, 44.6, and 45 years for the AC-T, TAC, and control groups respectively. Regarding the education level, about 17%, 34%, and 49% of the respondents had high school, diploma, and academic degrees respectively. None of the participants had a positive history of brain surgery, stroke, epilepsy, and other neurological disorders. Demographic characteristics of the participants are summarized in table 1.

**Table 1:** Demographic characteristics of the participants.

|  | *AC-T regimen*<br>*(n=16)* |  | *TAC regimen*<br>*(n=19)* |  | *Control group*<br>*(n=24)* |  |
| --- | --- | --- | --- | --- | --- | --- |
|  | No | % | No | % | No | % |
| ***Marital status*** |  |  |  |  |  |  |
| *Single* | 1 | 6.25 | 2 | 10.5 | 2 | 8.3 |
| *Married* | 13 | 81.25 | 15 | 79.0 | 22 | 91.7 |
| *Widowed* | 1 | 6.25 | 0 | 0 | 0 | 0 |
| *Divorced* | 1 | 6.25 | 2 | 10.5 | 0 | 0 |
| ***Education level*** |  |  |  |  |  |  |
| *Under diploma* | 2 | 12.5 | 3 | 15.8 | 3 | 12.5 |
| *Diploma* | 6 | 37.5 | 5 | 26.3 | 9 | 37.5 |
| *University education* | 8 | 50.0 | 11 | 57.9 | 12 | 50.0 |
| ***Occupation status*** |  |  |  |  |  |  |
| *Employed* | 3 | 18.75 | 4 | 21.0 | 6 | 25.0 |
| *Unemployed* | 12 | 75.0 | 15 | 79.0 | 16 | 66.7 |
| *Retired* | 1 | 6.25 | 0 | 0 | 2 | 8.3 |
| ***Time since the last chemotherapy regimen*** |  |  |  |  |  |  |
| *< 6 months* | 0 | 0 | 0 | 0 | - | - |
| *6-12 months* | 6 | 37.5 | 8 | 42.1 | - | - |
| *12-24 months* | 7 | 43.75 | 7 | 36.8 | - | - |
| *> 24 months* | 3 | 18.75 | 4 | 21.1 | - | - |

### 3.2 Hemoencephalography

To compare the cortical blood flow among the three groups, ANOVA test was used which showed that cortical hemodynamics had no significant difference not only in resting-state but also during task performance (p-value ≥ 0.05). Post-test cortical blood flow was shown to be significantly higher in the AC-T-treated breast cancer patients than in the control group (p-value = 0.02). The comparison between AC-T and TAC regimen and between TAC regimen and control group were not found to reveal significant differences (p-value ≥ 0.05). Moreover, repeated measures ANOVA test was used to assess the effect of task performance on cortical hemodynamics at FP1 which demonstrated increased blood flow during and after ANI task performance (control group: p-value= 0.009, F(2,46)=5.207; AC-T regimen: p-value=0.002, F(1.36,17.7)=11.353; TAC regimen: p-value=0.04, F(1.19,20.22)=4.519).

### 3.3. qEEG

The recorded qEEG files of each three groups were analyzed with the NeuroGuide software to generate the cortical color maps and statistical metrics. Since the purpose of this study was to evaluate attention deficits among chemotherapy-treated breast cancer patients and DAN and VAN networks comprise of fronto-parietal and centro-parietal regions, the fronto-centro-parietal electrodes including the F3, C3, P3, F4, C4, and P4 were selected for further analysis. The mean absolute power of these selected electrodes in beta1 and alpha frequency bands or sensorimotor rhythm (SMR) frequency is summarized in table 2. As the table indicates, the absolute power of both fronto-centro-parietal regions in SMR frequency is higher in the AC-T regimen than in the two other groups. Besides, the TAC regimen has higher absolute power in comparison to the control group. Figure 1 showed functional cortical maps representing the absolute power in beta 1 and alpha frequency bands in AC-T, TAC, and control groups during resting-state and ANI task performance.

**Table 2:**
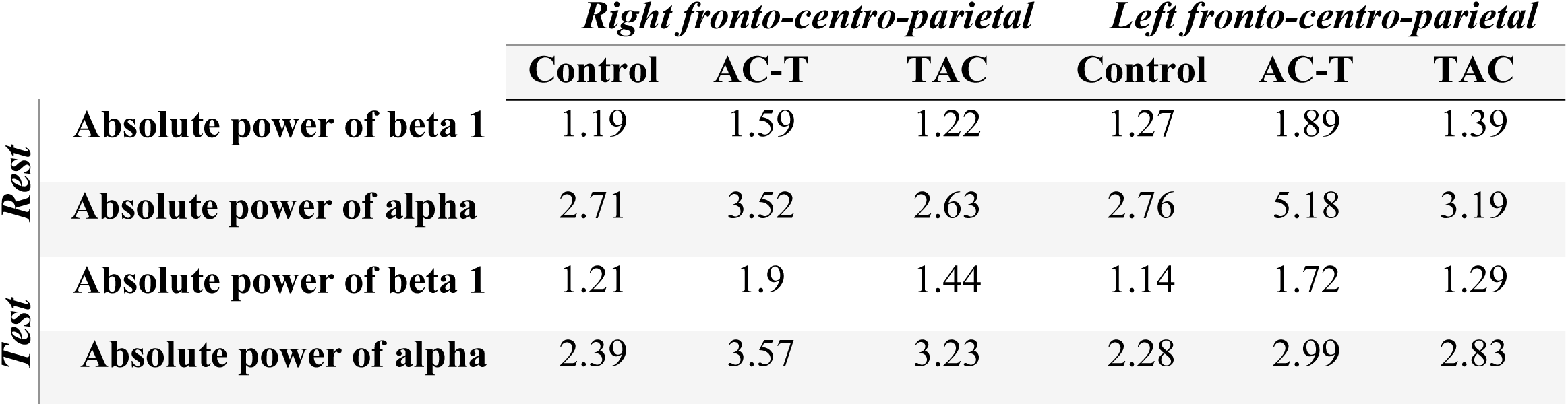
The mean absolute power of alpha and beta 1 frequency bands in fronto-centro-parietal electrodes in resting-state and task performance EEG data of AC-T, TAC, and control groups.

**Figure 1:**
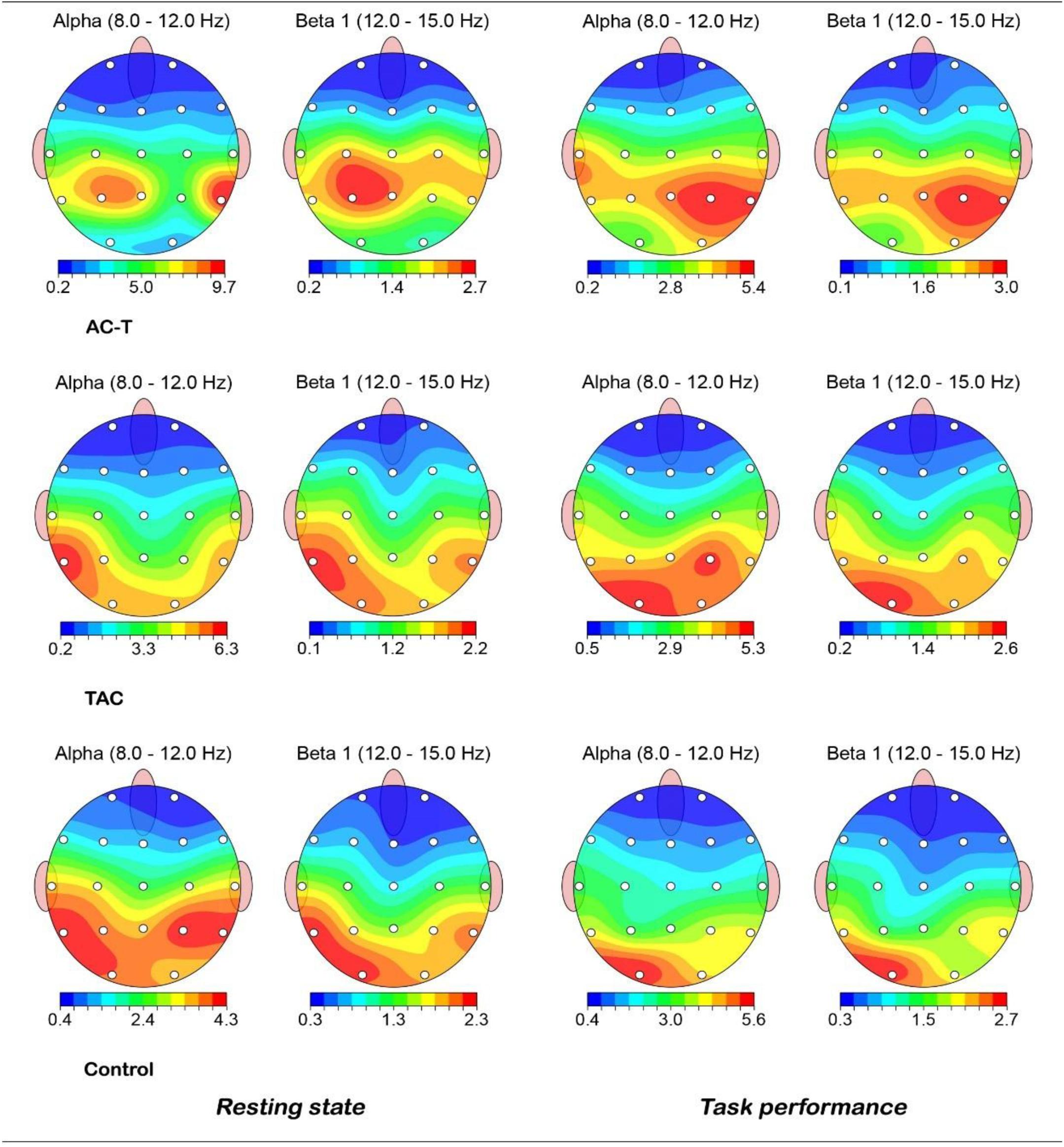
Absolute power in beta 1 and alpha frequency bands in AC-T, TAC, and control groups during resting-state and task performance.

To compare the mentioned groups, NGG analysis was conducted using the NeuroGuide software. Absolute power in the beta 1 frequency band in Cz and Pz electrodes was significantly higher in the AC-T regimen compared to the control group (p-value = 0.02 and 0.04 respectively). The AC-T group also had a significantly higher absolute power of the Cz electrode in the beta 1 frequency in comparison to the TAC regimen (p-value = 0.03). Moreover, absolute power in the FP1 electrode in beta 1 frequency was significantly higher in the control group than in the TAC regimen (p-value = 0.03) (figure 2). Other fronto-centro-parietal regions did not show significant differences.

**Figure 2:**
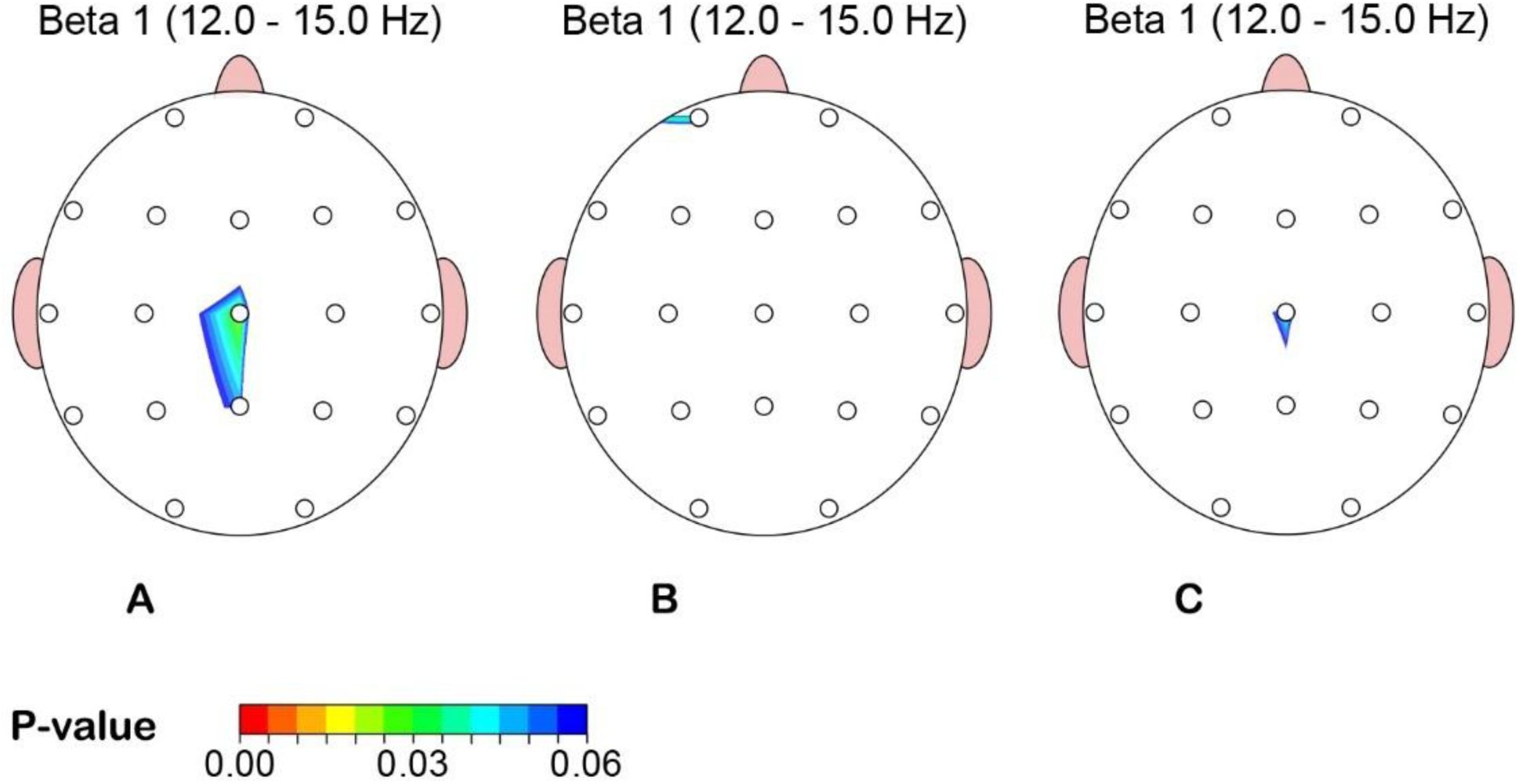
Cortical regions with significant differences in absolute power of beta 1 frequency band between AC-T regimen and control group (A), TAC regimen and control group (B), and AC-T vs TAC regimens (C).

Regarding the FFT coherence, fronto-centro-parietal electrodes with significant differences in the Z-scored FFT Coherence among the three groups (p-value ≤ 0.05) were analyzed. As it is shown in figure 3, the AC-T and TAC regimens had a decreased Z-scored coherence (hypocoherence) in the fronto-centro-parietal regions in SMR frequency compared to the control group. However, comparing the Z-scored coherence of the mentioned regions among the AC-T and TAC groups was inconclusive.

**Figure 3:**
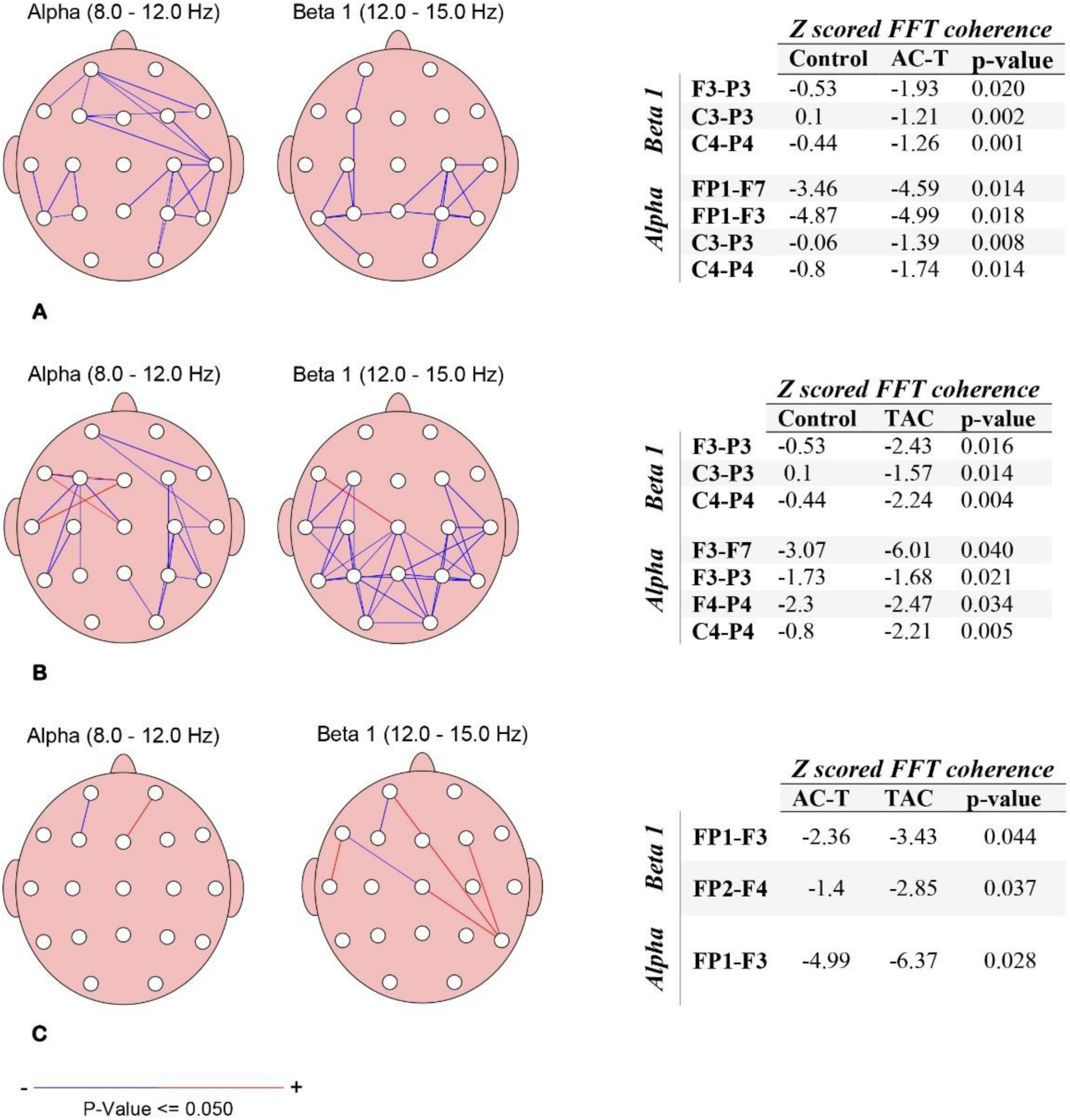
Cortical regions with significant differences in Z scored FFT coherence in alpha and beta frequency bands between AC-T regimen and control group (A), TAC regimen and control group (B), and AC-T and TAC regimens (C).

Assessment of CSD values of the DAN network centers in the control group using Neuronavigator analysis toolbox had shown that left Brodmann area 39 -left angular region-had the highest CSD value in both 10 and 15 Hz frequencies in resting-state and during task performance. On the other hand, among the VAN network nodes, left Brodmann area 21 - middle temporal region-had maximum CSD value in both 15 and 10 Hz in the control group (figure 4). As is illustrated in table 3, the CSD values were higher in the AC-T regimen than the two other groups in both DAN and VAN networks, while the control group had the lowest CSD values in all ROIs. Besides, these values decreased during task performance.

**Figure 4:**
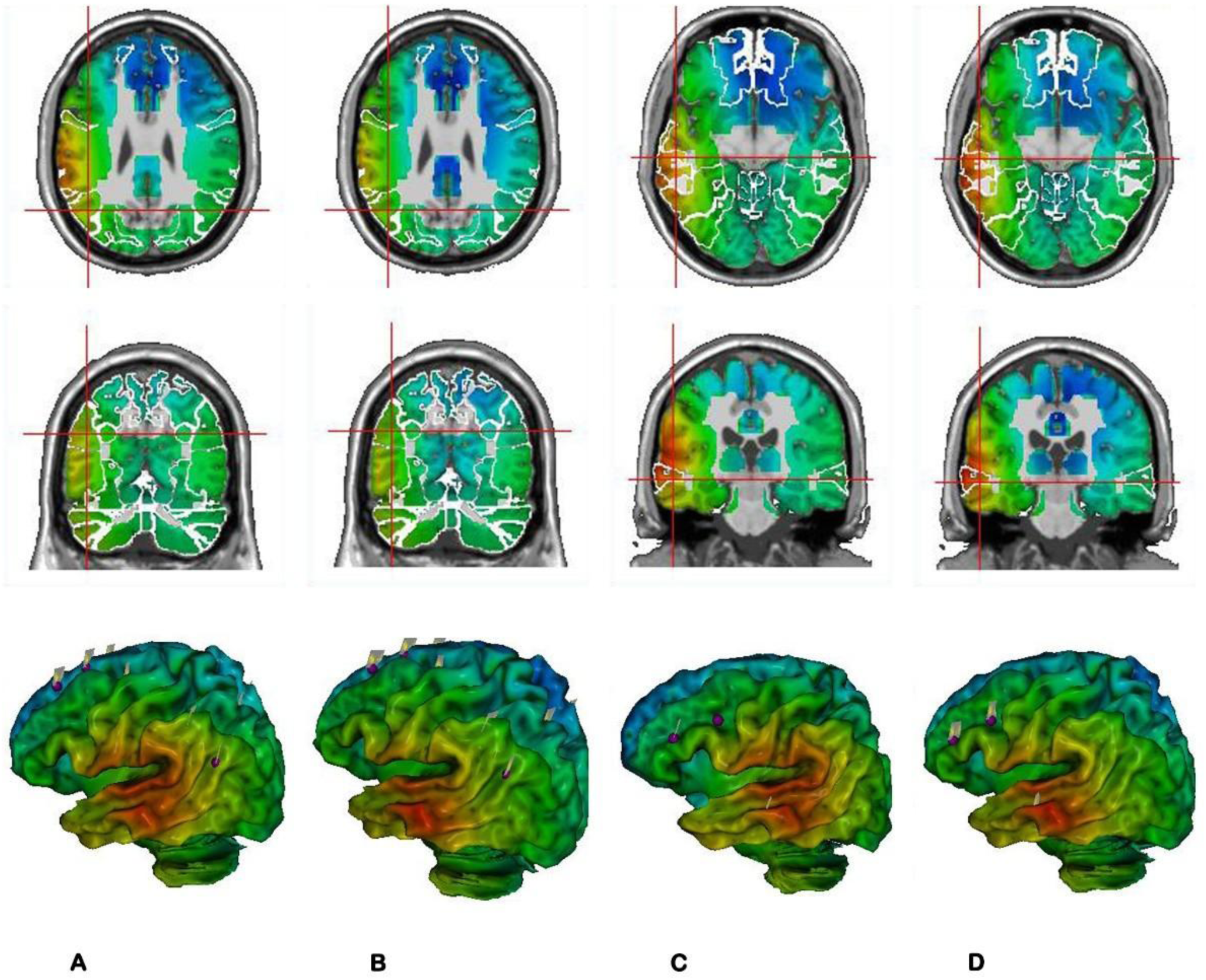
Coronal, sagittal and 3-D views of functional maps of current source density (CSD) values of brain regions in control group during resting-state with identification of centers involved in dorsal attention network (DAN) and ventral attention network (VAN) networks. Brain region with maximum CSD value is highlighted as maximum of the CSD values in 10 Hz frequency in DAN network (A), 15 Hz frequency in DAN network (B), 10 Hz frequency in VAN network (C), 15 Hz frequency in VAN network (D).

**Table 3:**
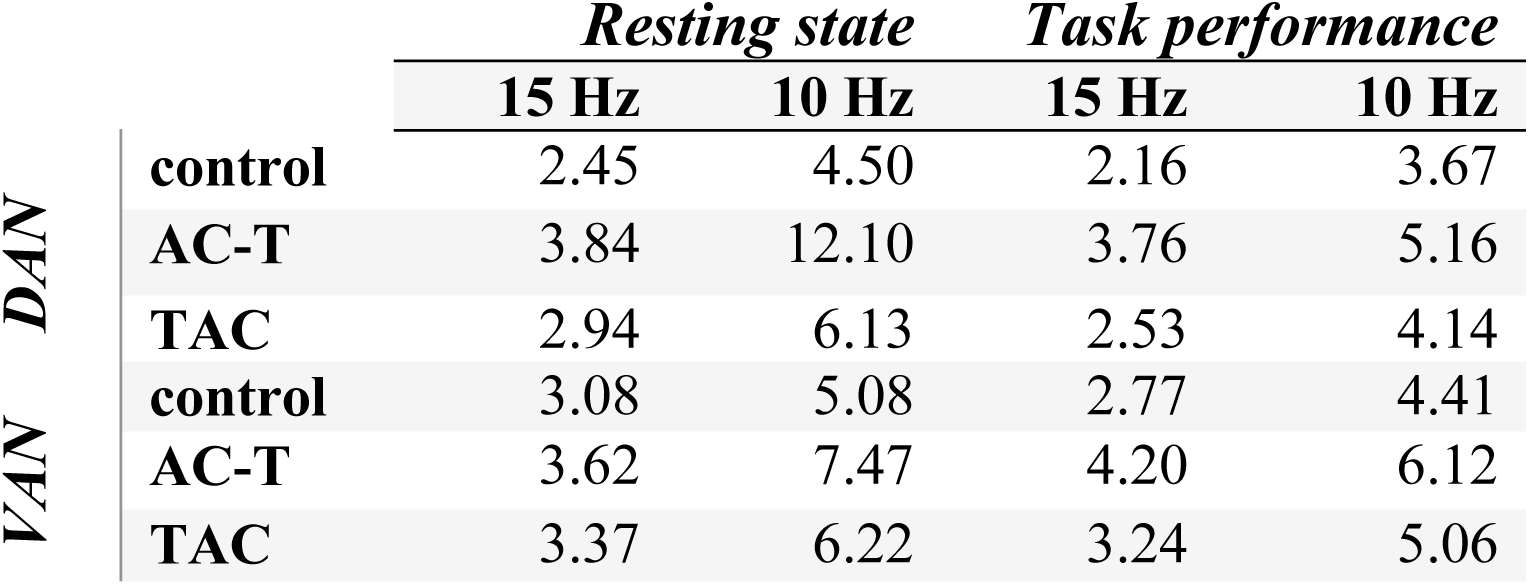
Maximum CSD values in the region of interests in 10 and 15 Hz frequencies in DAN and VAN networks.

## 4. Discussion

The theory of “chemobrain” is currently well established. However, very limited studies have evaluated the neuropsychological sequelae of different chemotherapy regimens to propose the least toxic regimen. Since the breast cancer patients receiving the AC-T regimen showed worse performance in sustained attention and visuospatial working memory which are regulated through the VAN and DAN networks, in the present study, we aimed to analyze functional cortical maps of fronto-parietal and centro-parietal areas in patients receiving AC-T and TAC chemotherapy regimens. The current study has revealed increased amplitude of alpha and beta 1 frequency bands in the AC-T regimen in comparison with the TAC regimen and control groups in these neural networks. In addition, the HEG study did not show significant differences in cortical hemodynamics between mentioned groups in both resting-state and test performance. However, post-test blood flow was shown to be higher in the AC-T regimen in comparison with the control group.

It is already known that stimuli processing includes multiple levels of hierarchical analysis named global (general features of subjects) versus local attentional processing (such as the cognitive tasks performed in the current study). It has been revealed that global and local attentional processing is lateralized in the brain; global processing is associated with the right hemisphere activation and local processing with the left hemisphere. Interestingly, this hemisphere asymmetry was also shown in the pre-stimulus resting intervals leading to better functional performance (40-42).

As observed, the mean absolute power of the beta 1 frequency band in the left fronto-centro-parietal regions was higher in the AC-T regimen than in the two other groups. Besides, the TAC regimen has higher absolute power in comparison with the control group. Moreover, the AC-T regimen had the highest CSD values in left Brodmann area 39 and left Brodmann area 21 in DAN and VAN networks in 15 Hz frequency. This pattern of left hemisphere activation is consistent with the hemisphere asymmetry seen in the local attentional processing.

Available literature has emphasized the important role of the beta 1 frequency band in cognitive function and particularly sustained attention (43, 44). For instance, a study published by Solís-Vivanco et al. showed that optimal attentional performance depends on the cooperation between DAN and VAN neural networks through modification of upper alpha/low beta (SMR) oscillations. They found a power increase of beta oscillations both in VAN and DAN nodes among the participants while performing a bimodal attention task (45).

Furthermore, increased beta power in the mentioned ROIs suggests greater cortical metabolic activity and elevated cognitive workload during task performance and is widely used for the measurement of mental workload while performing attentional tasks. Besides, individuals with higher beta activity at rest might have greater chronic cortical activation associated with psychological disorders such as obsessive-compulsive and generalized anxiety disorders (46-50).

Similarly, assessment of the mean absolute alpha band power in fronto-centro-parietal regions and CSD values in DAN and VAN networks in 10 Hz indicated the same results as beta 1 frequency band and 15 Hz, respectively. Regarding the alpha band, there are numerous studies evaluating its pattern in response to an increase in cognitive workload which have contributed to inconsistent results. Many of them indicated that a rise in mental workload is associated with the decrease of alpha and increase in theta band power (51-53). For Instance, in the study conducted by Mazher et al. to evaluate the cognitive workload during learning states, the alpha band had shown to have desynchronization with mental workload (54). MacLean’s study had the same results; in such a way that an increase in alpha and a decrease in beta are suggestive of less mental vigilance and attentional investment (55). On the other hand, some studies revealed augmentation of alpha band activation at the parietal region in response to an increase in cognitive load. They interpreted their findings with response inhibition theory which claims an increase in occipito-parietal alpha band power is associated with inhibition of irrelevant information (56, 57). Therefore, two distinct mechanisms can justify the synchronization and desynchronization of the alpha band. In line with this theory, in the current study, the AC-T regimen had higher alpha band power which can be indicative of more efforts for inhibition of irrelevant data and therefore increment of cognitive workload.

All the mentioned results reflect greater cognitive workload needed for the task performance, lack of cognitive ease, and potential development of post-chemotherapy cognitive impairment associated with some degrees of psychological disorders in AC-T-treated breast cancer survivors. Regarding the available literature, most of the studies evaluating chemobrain in breast cancer survivors used different chemotherapy protocols comprising different cytotoxic agents such as cyclophosphamide, epirubicin or doxorubicin, docetaxel or paclitaxel, doxorubicin, and 5-fluorouracil (58). However, most clinical research did not evaluate the relationship between different chemotherapy regimens and the associated cognitive impairment.

It is worth noting that the pathophysiology of chemotherapy-induced cognitive impairment is not well understood. McDonald et al. found a decrease in gray matter density in fronto-temporal regions in AC-T-treated breast cancer patients one month after chemotherapy with subsequent partial recovery after one year suggesting structural neuroanatomic causes for post-chemotherapy cognitive impairment. Although some possible etiologies including the chemotherapy-induced DNA damage and changes in neural plasticity can be considered, the exact mechanism of this gray matter decrease is unknown (59).

The AC-T regimen includes administration of doxorubicin (adriamycin) and cyclophosphamide (cytoxan) every three weeks for four cycles followed by paclitaxel (taxol) for four other cycles. Among these medications, taxane-induced neuropathy is well established. Paclitaxel administration causes microtubules aggregation in nerve axons leading to degenerative axonopathy. Besides the peripheral nervous system, the CNS axons are also at risk causing paclitaxel-induced encephalopathy (60-62). The same results were demonstrated in McElroy’s study in which the effects of the AC-T regimen in female mice were assessed. According to this study, oxidative stress resulting from the toxic effects of the AC-T regimen can lead to dendritic branching and morphologic alterations in the hippocampus besides the mitochondrial dysfunction which all contribute to cognitive dysfunction (63). In addition, since the agents of the AC-T regimen except cyclophosphamide cannot cross the brain blood barrier, one mechanism suggested for the chemobrain phenomenon is cytokine dysregulation and subsequent sustained inflammation induced by chemotherapeutic agents (64). The limited information regarding the pathophysiology of chemotherapy-induced cognitive decline highlights the need for conducting more detailed assessments.

The current study has some limitations. First, the baseline cognitive function of the participants before receiving the chemotherapy was not evaluated in this study due to the high levels of generalized anxiety and depressive disorders in the acute phase of the cancer diagnosis. Second, due to the COVID-19 pandemic and immunodeficiency of the breast cancer patients, the number of the cases enrolled in the study was limited to improve the safety of the participants. Besides, as Lange et al. mentioned, hormonal and targeted therapies in breast cancer patients might induce cognitive deficits (65). Although the results of available literature were inconclusive, recruiting exclusively chemotherapy-treated patients without a history of receiving hormonal therapies such as tamoxifen and aromatase inhibitors is reasonable to evaluate the impact of chemotherapy regimens. Therefore, more studies are needed to include a greater number of breast cancer patients treated with available chemotherapy regimens rather than the AC-T and TAC, two commonly prescribed regimens, to confirm the mentioned results and propose a therapeutic guideline for oncologists and other practitioners.

## Conclusion

This qEEG-based study revealed increased amplitude of high alpha/low beta (SMR) frequency bands in left fronto-centro-parietal regions in VAN and DAN networks suggestive of increased cognitive workload and a diminished cognitive function more specifically in the attention domain in breast cancer patients undergone AC-T chemotherapy regimen. The difference between the two chemotherapy regimens would potentially suggest a presumable toxic effect of the AC-T regimen on chemical dynamics of the brain which would result in changes in electro neurodynamics and ultimately cognitive attitude. This is what is commonly referred to as chemobrain.

## Data Availability

All data generated or analyzed during this study are available from the corresponding author on reasonable request.

## Acknowledgements

The authors would like to thank DANA Brain Health Institute; Iranian Neuroscience Society, Fars Chapter, Shiraz, Iran for the received support. The study received partial financial support from School of Advanced Medical Sciences and Technologies, Shiraz University of Medical Sciences, Shiraz, Iran (ID: 15307)

## Figures/Tables legends

**Table 1:** Demographic characteristics of the participants.

**Table 2:** The mean absolute power of alpha and beta 1 frequency bands in fronto-centro-parietal electrodes in resting-state and task performance electroencephalography (EEG) data of AC-T, TAC, and control groups.

**Table 3:** Maximum current source density (CSD) values in the region of interests in 10 and 15 Hz frequencies in dorsal attention network (DAN) and ventral attention network (VAN) networks.

